# Genome-wide association study of copy number variations in Parkinson’s disease

**DOI:** 10.1101/2024.08.21.24311915

**Authors:** Zied Landoulsi, Ashwin Ashok Kumar Sreelatha, Nicole Kuznetsov, Claudia Schulte, Dheeraj Reddy Bobbili, Ludovica Montanucci, Costin Leu, Lisa-Marie Niestroj, Emadeldin Hassanin, Cloé Domenighetti, Pierre-Emmanuel Sugier, Milena Radivojkov-Blagojevic, Peter Lichtner, Berta Portugal, Connor Edsall, Jens Kru□ger, Dena G Hernandez, Cornelis Blauwendraat, George D Mellick, Alexander Zimprich, Walter Pirker, Manuela Tan, Ekaterina Rogaeva, Anthony Lang, Sulev Koks, Pille Taba, Suzanne Lesage, Alexis Brice, Jean-Christophe Corvol, Marie-Christine Chartier-Harlin, Eugenie Mutez, Kathrin Brockmann, Angela B Deutschländer, Georges M Hadjigeorgiou, Efthimos Dardiotis, Leonidas Stefanis, Athina Maria Simitsi, Enza Maria Valente, Simona Petrucci, Letizia Straniero, Anna Zecchinelli, Gianni Pezzoli, Laura Brighina, Carlo Ferrarese, Grazia Annesi, Andrea Quattrone, Monica Gagliardi, Lena F Burbulla, Hirotaka Matsuo, Akiyoshi Nakayama, Nobutaka Hattori, Kenya Nishioka, Sun Ju Chung, Yun Joong Kim, Lukas Pavelka, Pierre Kolber, Bart PC van de Warrenburg, Bastiaan R Bloem, Andrew B. Singleton, Dan Vitale, Mathias Toft, Lasse Pihlstrom, Leonor Correia Guedes, Joaquim J Ferreira, Soraya Bardien, Jonathan Carr, Eduardo Tolosa, Mario Ezquerra, Pau Pastor, Karin Wirdefeldt, Nancy L Pedersen, Caroline Ran, Andrea C Belin, Andreas Puschmann, Carl E Clarke, Karen E Morrison, Dimitri Krainc, Matt J Farrer, Dennis Lal, the Global Parkinson Genetics Program (GP2), Alexis Elbaz, Thomas Gasser, Rejko Krüger, Manu Sharma, Patrick May, the Comprehensive Unbiased Risk Factor Assessment for Genetics and Environment in Parkinson’s Disease (COURAGE-PD) consortium

## Abstract

**Objective:** To investigate the impact of copy number variations (CNVs) on Parkinson’s disease (PD) pathogenesis using genome-wide data and explore their role in sporadic PD.

**Methods:** We analyzed CNV data from 11,035 PD patients (including 2,731 early-onset PD (EOPD)) and 8,901 controls from the COURAGE-PD consortium using a sliding window CNV-GWAS and genome-wide burden analysis. The independent dataset from the Global Parkinson Genetics Program (GP2) consisted of 23,089 cases and 18,824 controls were used to validate our initial findings.

**Results:** The exploratory dataset identifies multiple CNV regions associated with PD risk. The nominated CNV loci were not confirmed in an independent dataset, except that only a deletion in the *PRKN* gene, a well-established EOPD locus, remained genome-wide significant and robustly supported.

CNV burden analysis showed a higher prevalence of CNVs in PD-related genes in patients compared to controls (OR=1.56 [1.18-2.09], p=0.0013), with *PRKN* showing the highest burden (OR=1.47 [1.10-1.98], p=0.026). Patients with CNVs in *PRKN* had an earlier disease onset. Burden analysis with controls and EOPD patients showed similar results.

**Interpretation:** The largest CNV-based GWAS on PD highlights both the promise and pitfalls of array-based CNV detection in PD and underscores the relevance of whole-genome sequencing approaches in resolving the role of CNV in PD. The array-based findings are prone towards false positive findings that might arise either from platform limitations and/or cohort biases. Future studies require improved genotyping resolution and rigorous cross-cohort validation to reliably assess CNV contributions to PD risk.

## Introduction

A global survey estimates that Parkinson’s disease (PD) is the world’s fastest-growing neurodegenerative disorder, surpassing even Alzheimer’s disease ^1^. Concerted efforts are required to unravel the underlying complexity of a disease in which genetics and environmental factors appear to play an important role ^2,3^. To date, considerable progress has been made in understanding the genetic basis of PD by identifying families in which the disease segregates in Mendelian fashion and by applying array-based approaches (commonly used in genome-wide association studies (GWAS)) to identify risk factors for sporadic forms of PD ^4,5^.

GWAS have been successful in identifying a number of loci that are potentially relevant for PD ^6,7^. While this success has been remarkable, the genetic variability explained so far is between 19-39% ^6^ indicating that the genetic variability in the form of chromosomal arrangements - duplications, deletions - commonly referred to as copy number variations (CNVs) - may be an important player in explaining the genetic variability in PD ^8–11^. Indeed, previously studies identified several CNVs in PD-related genes. For example, genomic multiplications in the *SNCA* gene were shown to cause familial ^12^ and sporadic ^13^ forms of PD. Duplication or triplication of this gene can result in an excess of alpha-synuclein, which may then aggregate and form Lewy bodies, ultimately contributing to neurodegeneration observed in PD ^10^. Furthermore, CNVs in *PRKN* are the most prevalent CNVs among known PD genes ^14–16^, owing to its location in one of the most mutation-susceptible regions of the human genome ^17^. Deletions in familial PD forms were previously described in *PINK1* ^18,19^ and *PARK7* ^20,21^, but are less common than in *PRKN*. Unlike *SNCA*, deletions in homozygous-driven early-onset PD forms have demonstrated the role of loss of function genes in the etiology of the disease, while the pathogenic role of single heterozygous CNVs is still controversial.

The above examples support the role of CNVs in genes that are *bona-fide* loci for monogenetic PD. In contrast, the role and impact of CNVs on sporadic PD is still unclear. However, previous studies have suggested a potential role of CNVs in sporadic PD as well. For instance, a genome-wide CNV burden analysis in a Latin American cohort ^22^ showed that PD patients were significantly enriched only for CNVs affecting known PD genes, but they could not identify any putative CNV in PD pathogenesis at the genome-wide level. Their failure to identify putative CNVs could be attributed to the small sample size. Similarly, another study could only validate that the CNV within the *PRKN* locus was significantly associated with PD susceptibility ^23^. Thus, a CNV-based GWAS in large, well-powered and well characterized PD cohorts may reveal novel molecular pathways associated with the disease, potentially advancing efforts to understand the role of CNVs in PD pathogenesis.

The Comprehensive Unbiased Risk Factor Assessment for Genetics and Environment in Parkinson’s Disease (COURAGE-PD) is a worldwide collaboration consortium, aimed at understanding the roles of genetics and environment in PD ^7,24,25^. In the present study, we leveraged the genome-wide data to understand the impact of CNVs on PD in COURAGE-PD. Given the fact that there is no consensus on the best method for detecting or analyzing genome-wide significant CNVs ^9^, we applied the sliding window approach to perform CNV-GWAS, and employed genome-wide burden analyses to identify novel genome-wide significant CNV regions and to investigate their impact on the disease.

## Subjects and Methods

### Study cohort

The COURAGE-PD consortium includes data from 15,849 PD patients and 11,444 controls of predominantly European ancestry from 35 cohorts ^7^. In our study, we used genotyping data from 22,329 individuals from 25 European ancestry cohorts originating from 15 European countries. Genotyping quality control (QC) was conducted independently for each cohort, by following the standard procedure previously reported ^7^. Patients with early-onset PD (EOPD) were defined as those diagnosed before the age of 50 (age at onset (AAO) <= 50 years) ^26^. To replicate our findings, we used the NeuroBooster array data ^27^ from the large Global Parkinson Genetics Program (GP2) cohort comprising 23,089 PD patients and 18,824 controls.

### Copy number variant calling and quality control

We created a custom population B-allele frequency (BAF) and GC wave-adjusted log R ratio (LRR) intensity file using GenomeStudio (v2.0.5 Illumina) for all the samples that passed genotyping QC and employed PennCNV (v1.0.5, ^28^) to detect CNVs in our dataset. The analysis was restricted to autosomal CNVs, as calls from the sex chromosomes are often of poor quality ^28^. We used a post-CNV calling QC pipeline including standard parameters as previously described in studies on CNV calling from SNP array data for PD ^22^ or epilepsy ^29,30^. As a first step, adjacent CNV calls were merged into a single call if the number of overlapping markers between them was less than 20% of the total number when both segments were combined. This was followed by intensity-based QC to exclude samples with low-quality data. Samples with a LRR standard deviation of less than 0.24, an absolute value of the waviness factor (WF) of less than 0.03, and a BAF drift of less than 0.001 were retained. These thresholds correspond to the median plus 3 SDs. Furthermore, CNV calls with more than 50% overlap with known problematic genomic regions ^28^, including centromeric, telomeric, and HLA regions, were also excluded before analysis. Next, we removed CNVs that met the following criteria: they spanned fewer than 20 SNPs, were less than 20 kilobases (kb) in length, and had an SNP density of less than 0.0001 (number of SNPs/length of CNV). For CNVs spanning at least 20 SNPs and longer than 1 Mb, SNP density was not considered. Finally, we applied a series of filters to identify rare CNVs. The first step was to assign a specific frequency count to each CNV call using PLINK v.1.07 ^31^. This was followed by applying a filter to exclude common CNVs, retaining only rare variants that overlapped with CNVs in at least 1% of all samples. In the second step, CNVs with an overlap of at least 50% with reported common CNVs (allele frequency >1%) in the Database of Genomic Variants (DGV Gold standard dataset ^32^) and DECIPHER population CNVs frequencies ^33^were excluded. The filtered rare CNVs were annotated for gene content using refGene, including the gene name and corresponding coordinates in the hg19 assembly with ANNOVAR (v 2020-06-08).

### Sliding windows CNV analysis and assessment of genome-wide significance

A segment-based rare CNV burden analysis was performed to identify genomic regions with a significant increase in rare CNVs in PD cases compared to controls. This analysis was conducted separately for each type of CNV (deletion or duplication) using a sliding window approach ^34^. The sliding windows model allowed association testing of all autosomes through 267,237 sliding windows characterized by a window size of 200 kb and a step size of 10 kb, corresponding to 13,339.6 non-overlapping windows. The threshold for genome-wide significance was set to α =3.74 × 10e-06 after Bonferroni correction for multiple testing. For each of the genomic regions, the number of overlapping CNVs was counted separately for cases and controls for deletions or duplications. We considered a minimum overlap of 10% between the CNV and the genomic window to identify the potential burden of small deletions or duplications (≥ 20 kb). We used two statistical approaches to evaluate genome-wide significant windows. First, we applied a one-sided Fisher’s exact test, following the original method ^34^. Second, we performed logistic regression, modelling disease status as the dependent variable and CNV status as the primary independent variable. The regression model was adjusted for sex, age, sub-cohort, and the first five principal components from the population stratification procedure to account for potential confounding factors. The analysis was performed using the rCNV docker (https://hub.docker.com/r/talkowski/rcnv) ^34^, and custom Python (version 3.7.9) and R (version 4.3.1) scripts. To assess the impact of age at PD onset, the same analysis was subsequently stratified based on EOPD, encompassing all the control subjects and patients with EOPD.

### Association fine-mapping

The observation that a considerable number of large rare CNVs involve the deletion or duplication of multiple adjacent genes, led us to hypothesize that the most associated genes were not causal, but rather gained significance due to proximity to true causal genes. This is analogous to the linkage disequilibrium effect observed in GWAS of common variants. To address this issue, Collins *et al*. employed a Bayesian fine-mapping algorithm to define the 95% credible set of causal elements or genes at each genome-wide significant locus for deletions and duplications ^34^. This method prioritizes the most probable causal genes based on their association statistics. The Bayesian algorithm was employed to calculate the approximate Bayes factor (ABF) for each window, as previously described by Wakefield ^35^. The ABF offers an alternative to the P-values for assessing the significance of association by providing a summary measure that ranks these associations. Bayesian model averaging was used to estimate the null variance of true causal loci across taking into account prior information regarding the mean of all significant windows and the most significant window per block. The minimal set of windows that constitutes the 95% credible set for each block was defined by ranking the windows in descending order according to their ABF.

### CNV replication using CNV-Finder

To replicate CNVs identified in our discovery dataset, we applied CNV-Finder (https://github.com/GP2code/CNV-Finder) ^36^, a deep learning-based pipeline designed for CNV detection from Illumina genotyping arrays. CNV-Finder utilizes a Long Short-Term Memory (LSTM) neural network trained on expert-annotated LRR and BAF signals to detect deletions and duplications within predefined genomic regions. The pipeline allowed for filtering based on prediction confidence, LRR signal range, and variant count. Predictions were reviewed visually and used to prioritize samples for downstream validation.

### Genome-wide burden and survival analysis

The burden of rare CNVs associated with PD was calculated using distinct categories to determine their relative impact on PD risk, as previously reported ^22^. These categories included: (1) carrier status of genome-wide CNV burden, including CNVs in non-genic regions, for all CNVs, not distinguishing between deletions and duplications (2) carrier status of any exonic or intronic CNVs intersecting with ‘any gene’ except those associated with PD, (3) carrier status of CNVs intersecting with exonic or intronic regions of the six major ‘PD-related genes’ according to MDS gene classification (https://www.mdsgene.org): *LRRK2, SNCA, VPS35* for dominant forms of classical parkinsonism and *PRKN, PARK7*, and *PINK1* for recessive forms of early-onset parkinsonism, (4) carrier status of CNVs intersecting only with exonic regions of the ‘PD-related genes’, (5) carrier status of CNVs in *PRKN* only and (6) carrier status of large CNVs (≥1 Mb in length). To compare the CNV burden between PD patients and controls, we used the *glm* function in R (v4.3.1) to fit a logistic regression model. This allowed the calculation of the OR with a 95% confidence interval and p-values. We used sex, age at assessment, and the first five principal components (PCs) from the population stratification as covariates for the regression model. Cox proportional hazards regression analyses and Kaplan– Meier curves were generated using the *survival* R package ^37^ with age defined as age at last visit for controls and AAO for cases. Controls were included as censored observations given that it was only known that they did not develop PD up to their last visit. Hazard ratios (HRs) and 95% confidence intervals (CIs) of PD were estimated by Cox proportional hazards models. Sex and the first five PCs were included as covariates. All the P-values underwent adjustment using the Bonferroni method to correct for multiple testing.

## Results

### COURAGE-PD cohort

The final dataset for the COURAGE-PD cohort comprised 11,035 patients with PD and 8,901 control, all of European descent, following the QC steps for the genotyping data. The demographic characteristics of each cohort are presented in Supplementary Table 1. A total of 2,731 PD patients (24.7% of the total patients) were identified as having EOPD (mean AAO 43.1±6.1 and mean age at assessment 54.4±9.3 years). A schematic overview of the study design and analytical workflow is presented in Figure 1. We initially detected 1,098,221 CNVs in a total of 10,877 PD patients and 8,534 controls. After all QC and filtering steps (see Methods), the final number of rare CNVs was 28,263, including 7,843 duplications and 20,420 deletions in 3,896 PD patients and 3,299 controls. CNV analysis showed that 36.0% of the samples carried at least one QC-passed CNV. The characteristics of our CNV analysis are shown in Supplementary Table 2.

**Figure 1:**
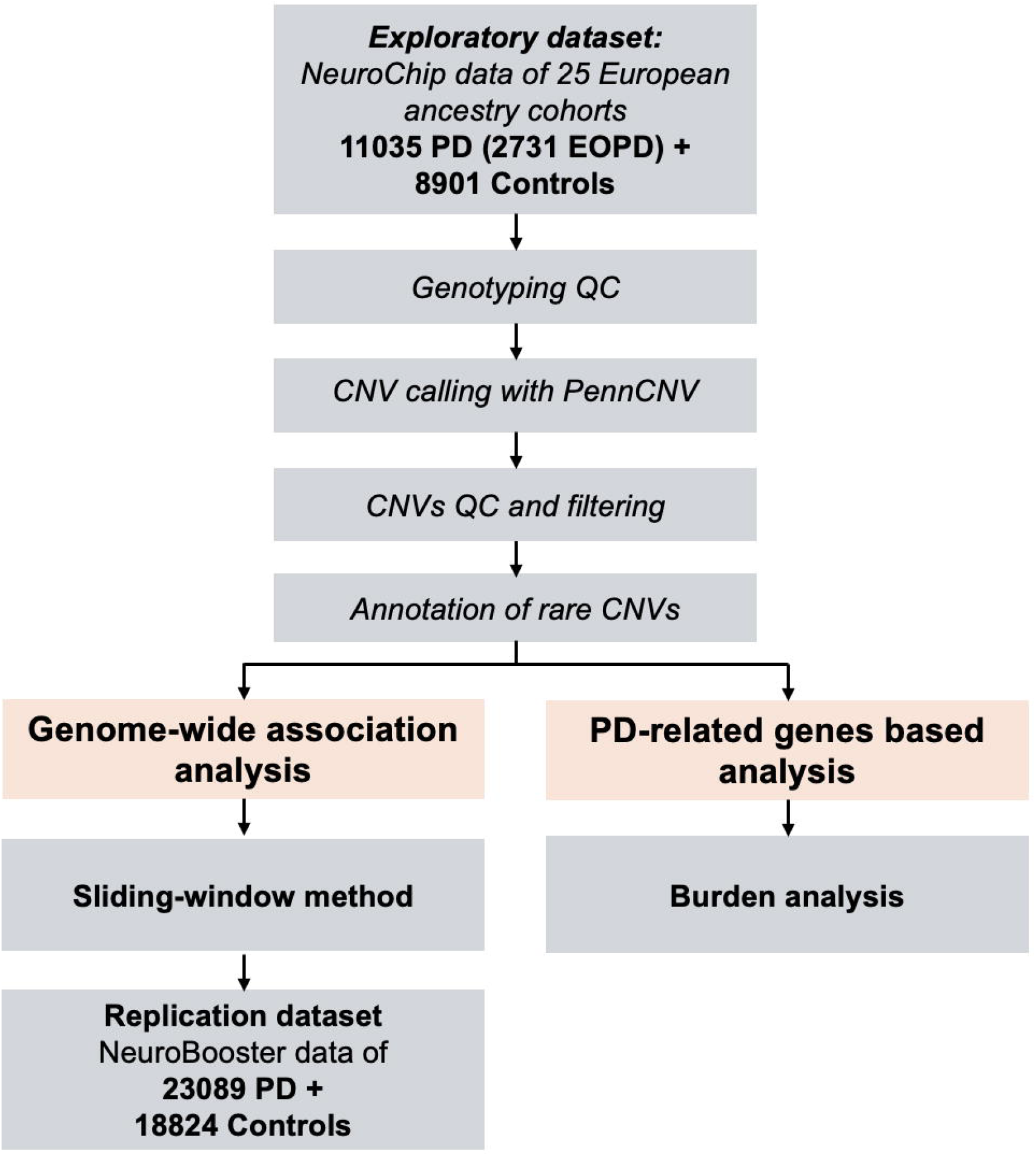
Overview of the study: schematic representation of the analysis workflow Study design.

### CNVs GWAS using sliding window approach with Fisher’s exact test: Identification of 14 genome-wide significant PD-associated CNV regions

A total of 267,237 genomic segments of 200 kb size in a 10 kb sliding window approach were scanned ^34^. After adjusting for the multiple testing and applying the fine-mapping approach, we identified 14 genome-wide significant CNV loci associated with PD. The 14 loci comprised one deletion (210 kb) and 13 duplications (size range: 220 to 560 kb, Fig 2A and Table 1). The genome-wide deletion identified in this study was a single-copy loss and spanned two genes *COL18A1, and POFUT2* (Table 1). All duplications, except for the two-copy gain in 8q23.3, were single-copy gains. They spanned a total of 68 genes, with three duplications in 7q22.1, 11q12.3, and 7q33 showing the highest ORs (Table 1).

**Figure 2:**
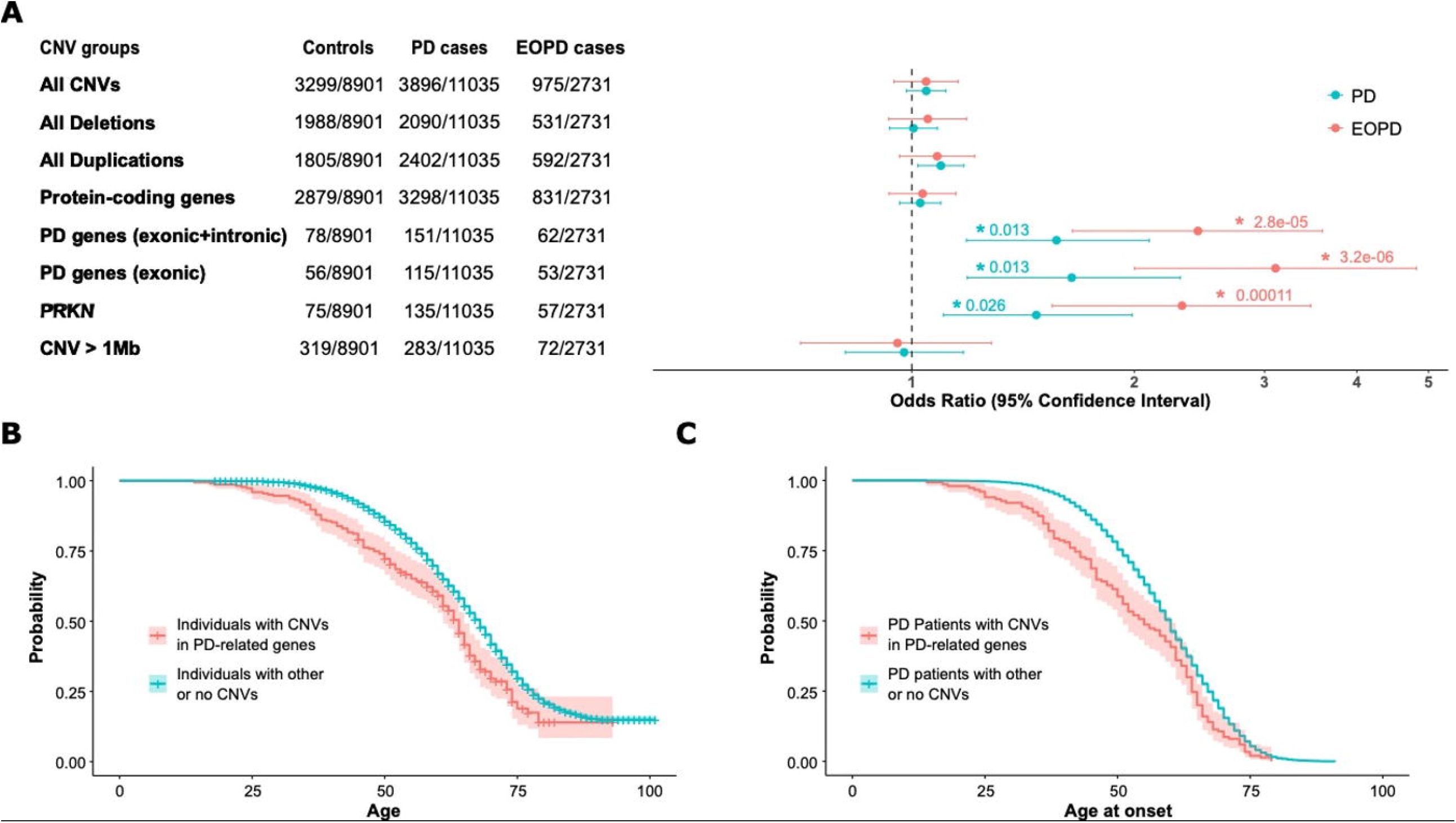
Genome-wide meta-analysis of PD. Miami plot of the CNV genome-wide association analyses illustrating the -log10 transformed Bonferroni-corrected p-values (DEL and DUP for deletions at the top and duplications at the bottom, mirrored respectively) for the enrichment of CNVs in cases vs. controls for each 200 kb sliding window. Results are shown for Fisher’s exact tests (A) and logistic regression before (B) and after (C) the exclusion of 220 samples from the Tübingen cohort carrying genome-wide significant CNVs, Adjacent chromosomes are shown in alternating light and dark colors. Genomic regions that exceeded the Bonferroni-corrected significance threshold (blue line, α =3.74×10^−6^) were annotated with the genomic band containing the signal.

**Table 1:** Genome-wide significant CNV regions in Parkinson’s disease. Column 1: Cytoband localization of the CNV. Column 2: Credible interval showing the genomic coordinates (start and end position) of the credible CNVs region supported by genome-wide association signals containing the causal element/gene with 95%. Column 3: CNV types with DEL and DUP indicate deletions and duplications. Columns 4 and 5: the lowest p-values in each CNV region and corresponding odds ratio (OR) with a 95% confidence interval. Columns 6 and 7: list of genes overlapping the genome-wide significant CNVs regions and known disease genes in these regions with causal genes between brackets.

| Cytoband | Credible interval | CNV type | Lowest p-value (neg log<sub>10</sub> p) | OR [95% CI] | CNV overlapping genes | Known disease genes |
| --- | --- | --- | --- | --- | --- | --- |
| 1q42.12 | chr1:224960000-225410000 | DUP | 23.85 | 21.18 [8.24-54.42] | *DNAH14* | Neurodevelopmental disorders (*DNAH14*) |
| 4q22.1 | chr4:90700000-90930000 | DUP | 5.47 | 7.68 [2.55-23.18] | *MMRN1*, *SNCA* | Parkinson's disease (*SNCA*) |
| 6p21.2 | chr6:38620000-39000000 | DUP | 5.8 | 5.98 [2.45-14.63] | *DNAH8*, *GLO1* | Spermatogenic failure (*DNAH8*) |
| 6q21 | chr6:109680000-110220000 | DUP | 18.9 | 13.01 [5.87-28.81] | *AK9*, *CD164*, *FIG4*, *MICAL1*, *PPIL6*, *SMPD2*, *ZBTB24* | Charcot-Marie-Tooth disease type 4J (*FIG4*), Amyotrophic lateral sclerosis type 11 (*FIG4*) |
| 7q22.1 | chr7:99080000-99560000 | DUP | 7.48 | 48.62 [2.97-796.61] | *CYP3A4*, *CYP3A43*, *CYP3A5*, *CYP3A7*, *CYP3A7-CYP3A51P*, *FAM200A*, *GJC3*, *OR2AE1*, *TMEM225B*, *TRIM4*, *ZKSCAN5*, *ZNF394*, *ZNF655*, *ZNF789*, *ZSCAN25* | - |
| 7q33 | chr7:134750000-134980000 | DUP | 6.41 | 41.75 [2.54-686.79] | *AGBL3*, *CYREN*, *STRA8*, *TMEM140*, *WDR91* | - |
| 8q23.3 | chr8:113780000-114340000 | DUP | 8.45 | 3.24 [2.11-4.99] | *CSMD3* | - |
| 11q12.3 | chr11:62910000-63350000 | DUP | 7.48 | 48.62 [2.97-796.61] | *LGALS12*, *PLAAT2*, *PLAAT3*, *PLAAT4*, *PLAAT5*, *SLC22A10*, *SLC22A24*, *SLC22A25*, *SLC22A9* | - |
| 12q24.12 | chr12:111790000-112010000 | DUP | 5.97 | 3.46 [1.98-6.04] | *ATXN2*, *PHETA1*, *SH2B3* | Spinocerebellar ataxia type 2 (*ATXN2*), Susceptibility to: Amyotrophic lateral sclerosis type 13 (*ATXN2*) and Late-onset Parkinson's disease (*ATXN2*), |
| 13q22.3 | chr13:77490000-77780000 | DUP | 5.73 | 6.74 [2.53-17.98] | *ACOD1, CLN5, FBXL3, MYCBP2* | Neuronal ceroid lipofuscinosis (*CLN5*), Intellectual developmental disorder with short stature, facial anomalies, and speech defects (*FBXL3*) |
| 15q22.2 | chr15:62070000-62360000 | DUP | 7.95 | 5.03 [2.61-9.69] | *C2CD4A, VPS13C* | Parkinson's disease (*VPS13C*) |
| 19p13.11 | chr19:17580000-17960000 | DUP | 6.64 | 7.5 [2.83-19.91] | *B3GNT3, COLGALT1, FCHO1, INSL3, JAK3, MAP1S, NIBAN3, PGLS, SLC27A1, UNC13A* | Immunodeficiency 76 (*FCHO1*), T-B+ severe combined immunodeficiency (*JAK3*) Brain small vessel disease 3 (*COLGALT1*) |
| 20p13 | chr20:3380000-3740000 | DUP | 7.81 | 3.43 [2.13-5.51] | *ADAM33, ATRN, DNAAF9, ADISSP, GFRA4, HSPA12B, SIGLEC1* | - |
| 21q22.3 | chr21:46650000-46860000 | DEL | 6.64 | 3.46 [2.05-5.85] | *COL18A1, POFUT2* | Knobloch syndrome type 1 (*COL18A1*), Glaucoma, primary closed-angle (*COL18A1*) |

Of the 14 genome-wide significant CNV regions identified in this study, none overlapped with the most recent SNP-based GWAS study ^6^. However, two genome-wide significant CNV regions overlapped with two PD-related genes, namely *SNCA* and *VPS13C* (Table 1). The 4q22.1 duplication interval encompasses exons 1 to 4 of *SNCA* and the 15q22.2 duplication interval overlapped with the entire coding region of *VPS13C*. These two rare duplication events did not overlap with the common significant GWAS SNPs identified in *SNCA* and *VPS13C* ^6^. Furthermore, the correlation between these SNPs and the closest duplication SNP demonstrated that these loci are in complete linkage equilibrium (r^2^ = 0).

### Assessment of significant CNV regions in the Global Parkinson’s Genetics Program (GP2) dataset

To replicate our findings, we utilized the CNV-Finder pipeline ^36^ applied to NeuroBooster array ^27^ data from the large GP2 cohort comprising 23,089 PD patients and 18,824 controls. None of the previously identified regions were visually confirmed, nor did they occur frequently enough to account for a significant enrichment in PD compared to controls indicating the initial novel CNV-loci are false-positive signals

### Evaluating cohort-specific signals and functional validation

To explore potential reasons for the lack of replication in the GP2 dataset, we reanalyzed our data using the same sliding window approach and applied logistic regression to adjust for multiple potential confounders, including sub-cohort heterogeneity. This analysis identified three duplication regions of genome-wide significance (1q42.12, 6q21, and 19p13.11; see Fig 2B) that remained significant after correction for multiple testing. As expected, these regions corresponded to the top signals previously observed in the CNV GWAS based on Fisher’s exact test.

We screened a subset of samples from one COURAGE-PD site (Gasser/Sharma study; see Supplementary Table 1) using the multiplex ligation-dependent probe amplification (MLPA) assay. Of the 12 PD patients tested for 1q42.12, nine for 6q21, and nine for 19p13.11, none showed evidence of a duplication or deletion within the corresponding regions.

Upon closer examination of CNV carrier distribution across the genome-wide significant regions identified through Fisher’s exact test and/or logistic regression, we observed a striking overrepresentation of samples from a single sub-cohort, namely the Gasser/Sharma study from Tuebingen (see Supplementary Table 1). In several regions, between 50% and 80% of CNV carriers originated from this sub-cohort, despite it representing only about 15% of the total study population. This cohort-specific inflation was not detectable through standard covariate adjustment, using principal components and cohort indicator variables, suggesting the presence of technical or biological artefacts specific to this sub-cohort that warrant further investigation.

The cohort-specific nature of the signals only became evident after we performed leave-one-out GWAS analyses, sequentially excluding one sub-cohort at a time. These analyses revealed that the strongest genome-wide signals were almost entirely driven by samples from the Tuebingen sub-cohort (Supplementary Figure 1). Notably, this was also the sub-cohort from which individuals had been selected for molecular validation; for none of these individuals the reported duplications could be validated. Excluding the 220 samples carrying genome-wide significant CNVs from this sub-cohort resulted in the loss of all genome-wide significant signals (Fig 2C), indicating that the associations were likely cohort-specific artefacts rather than true biological findings. The same findings were observed even after removing the entire sub-cohort (data not shown). Moreover, carriers within this sub-cohort did not form a distinct principal components cluster. The significant CNVs identified in these samples, were also not identified as outliers based on standard CNV signal quality metrics (including LRR, BAF or WF), nor were detected in regions with particularly low SNPs coverage.

### Impact of SNP Density on CNV Association Signals and Cohort-Specific Inflation

The Neurochip array used for CNV detection includes a common variant backbone (HumanCore-24, Illumina) comprising approximately 306k SNPs, supplemented by ∼180k neurological disease-specific markers ^38^. To investigate the cohort-specific inflation observed in our CNV-GWAS results, we reevaluated the association using only the common variant backbone, applying the sliding window approach. After multiple testing correction and fine-mapping, we identified six genome-wide significant CNV loci associated with PD (Supplementary Figure 2.A). However, these loci were no longer significant when the Tuebingen cohort was excluded from the analysis, replicating the inflation pattern observed with the full Neurochip array (Supplementary Figure 2.B). This underscores the disproportionate influence of this single cohort on the association results.

### Identification and confirmation of a PRKN deletion region in EOPD

To gain further insight into the genome-wide landscape of EOPD, we conducted a subset analysis comparing 2,731 PD patients with an AAO less than 50 years old and controls. This analysis identified four loci reaching genome-wide significance (Figure 3A). Three of these regions (1q42.12, 6q21, and 11q12.3) corresponded to duplications previously flagged as likely false positives in the GWAS of the full cohort (Figure 2A and Table 1). In contrast, one novel deletion region spanning ∼590 kb on chromosome 6q26 (chr6:162,340,000–162,930,000) emerged as a robust signal. This region overlaps exons 2–6 of *PRKN*, a gene well known for its association with autosomal recessive EOPD, with an odds ratio of 6.8[3.52–13.14]. Even after removing the 220 CNV carriers from the Tübingen cohort, which exhibited a cohort-specific imbalance, the *PRKN* deletion retained genome-wide significance (Figure 3B). To validate these findings, we screened CNVs within this region in the Tübingen cohort using MLPA. Among 26 individuals with *PRKN* CNVs (21 patients and 5 controls), MLPA confirmed the presence of deletions in 22 individuals, yielding an overall confirmation rate of 85%. Replication using the CNV-Finder pipeline in the independent GP2 dataset further validated the *PRKN* deletion signal, demonstrating a significant enrichment of carriers among PD patients compared to controls (OR = 1.5[1.18-1.93], p = 1.6e-05).

**Figure 3:**
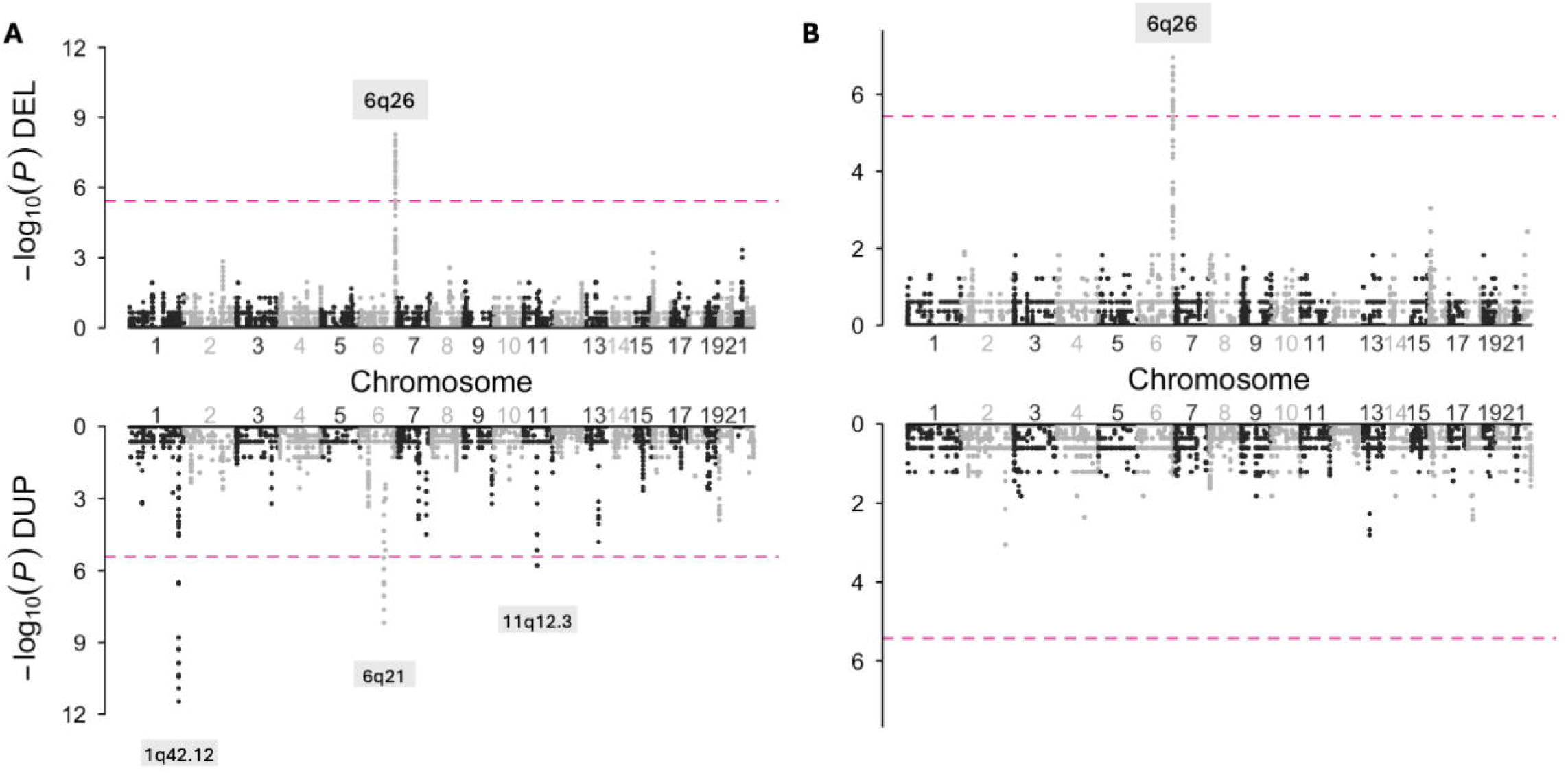
Genome-wide meta-analysis of EOPD. Miami plot of the CNV genome-wide association analyses illustrating the -log10 transformed Bonferroni-corrected p-values (DEL and DUP for deletions at the top and duplications at the bottom, mirrored respectively) of the Fisher’s exact tests for the enrichment of CNVs in cases vs. controls for each 200 kb sliding window before (A) and after (B) the exclusion of 220 samples from the Tübingen cohort carrying genome-wide significant CNVs. Adjacent chromosomes are shown in alternating light and dark colors. Genomic regions that exceeded the Bonferroni-corrected significance threshold (blue line, α =3.74×10^−6^) were annotated with the genomic band containing the signal.

### CNV burden analysis

We used logistic regression to compare the rare CNV burden on a genome-wide level in PD patients and controls across the predefined categories (see Methods). No significant difference was observed for genome-wide CNV burden (OR=1.04 [0.98-1.11], adjusted p-value (p_adj_)=0.3), duplications (OR=1.09 [1.01-1.17], p_adj_ = 0.46), or deletions (OR=1.00 [0.93-1.08], p_adj_=0.89, Fig 4A), implicating that cumulative burden of CNVs does not alter the risk of getting PD. The same results were observed for the CNVs burden for all the genes excluding PD-related ones (OR=1.02 [0.96-1.09], p_adj_=0.56) and large CNVs exceeding 1Mb in length (OR=0.97 [0.81-1.17], p_adj_=0.89, Fig 4A).

**Figure 4:**
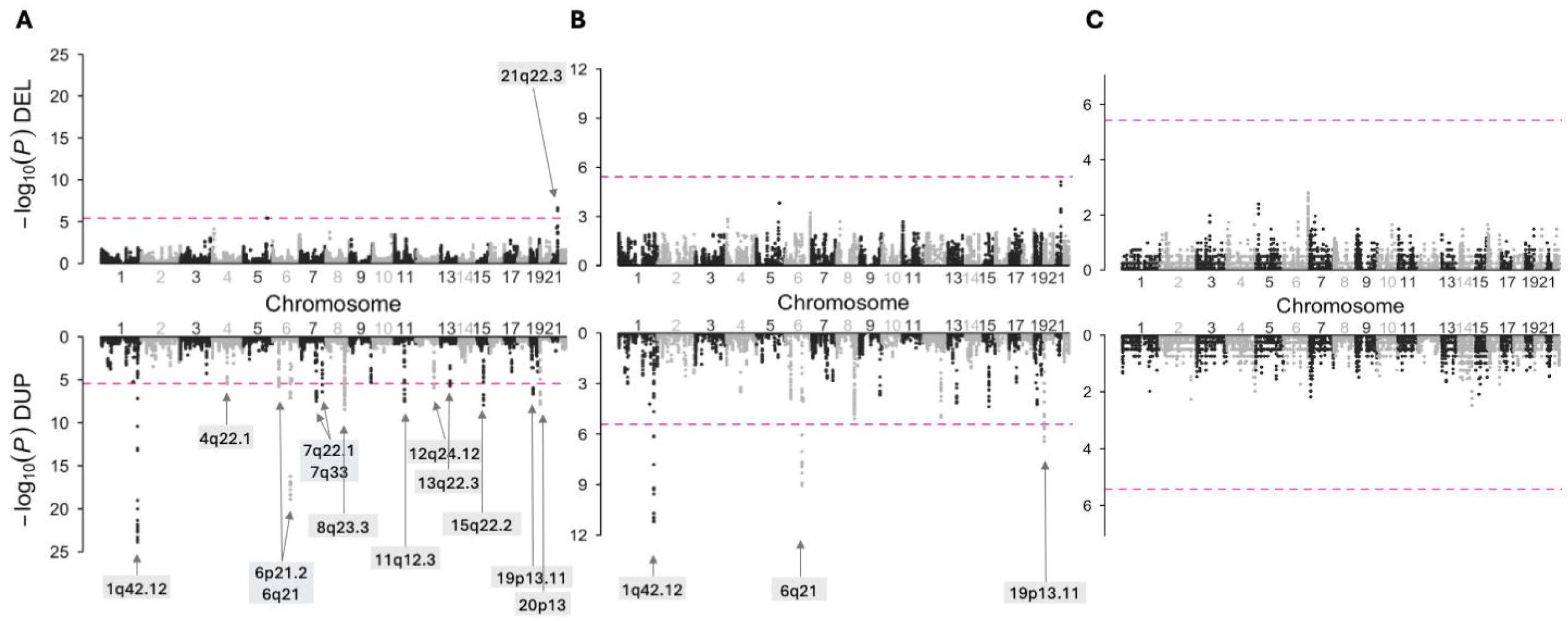
Rare CNV burden in PD and Early-onset PD patients compared to controls for different categories. **A**. Logistic regression was used to calculate odds ratios (ORs) and Bonferroni-adjusted p-values for each CNV category, and were adjusted for age, sex, and the first five components of PCAs. Protein-coding gene categories were defined as all coding genes except PD-related genes. * Bonferroni adjusted p-values surpassing the multiple testing cut-off. **B-C**. Kaplan– Meier estimates of individuals (PD patients and controls in **B** and PD patients only in **C**) carrying a CNV in a PD-related gene and individuals with other or no CNVs. Probability: the probability of not having PD symptoms. Age: age at last visit for controls or age at onset for cases. Highlighting around curves indicates 95% confidence intervals.

In contrast, we observed a significant CNV burden for PD-related genes when considering all CNVs (OR=1.56 [1.18-2.09], p_adj_=0.0013) with a slightly more pronounced association for CNVs overlapping exonic regions (OR=1.64 [1.18-2.30], p_adj_=0.013, Fig 4A). CNVs in *PRKN* were the primary contributors to this finding (OR=1.47 [1.10-1.98], p_adj_=0.026, Fig 4A). Indeed, among the 229 individuals carrying CNVs in PD-related genes, 91.7% were carriers of *PRKN* CNVs (summarised in Supplementary Table 3 and detailed in Supplementary Table 4). All these *PRKN* CNVs mostly consisted of one-copy duplications or duplications, except for seven homozygous deletions in PD patients (Table S3-4). CNVs in *PRKN* were identified in 135 PD patients (115 exonic and 36 intronic, representing 1.2% of the total patients) and 75 controls (56 exonic and 22 intronic, representing 0.8% of controls, Table S3-4). The AAO for patients with CNVs in *PRKN* was significantly earlier than that of patients without CNVs in *PRKN* (52.8±11.5 vs. 58.7±15.0 years, Mann-Whitney test p-value = 2.5e-05). Moreover, patients carrying exonic *PRKN* variants exhibited a significantly earlier AAO (50.8±15.5 years) compared to those carrying intronic variants (58.0 ± 12.2 years, Mann-Whitney test p-value = 0.02).

### EOPD CNV burden analysis

We also performed a subset analysis comparing only the 2,731 cases with AAO ≤ 50 years with controls to evaluate the rare CNV burden in patients with EOPD. Again, our findings revealed a significant increase in CNVs in PD-related genes (*OR*=2.43 [1.64-3.59], *p*_*adj*_=2.8e-05) with a slightly stronger association when examining CNVs that intersected with exonic regions (*OR*=3.10 [2.00-4.81], *p*_*adj*_=3.2e-06, Fig 4A). Among the EOPD cohort, 62 patients (2.2% of EOPD patients) were identified as carriers of CNVs in PD-related genes, with *PRKN* being the most common (*OR*=2.32 [1.54-3.46], *p-value*_*adj*_=1.1e-04, Fig 4A), found in 57 patients (2.1% of EOPD patients), encompassing 48 exonic and seven intronic CNVs. Carriers of the seven homozygous *PRKN* deletions mentioned above were EOPD patients.

To further investigate the role of CNVs in PD subtypes, we compared EOPD cases to LOPD cases. This analysis revealed a significantly increased CNV burden in EOPD patients compared to LOPD patients, mirroring the trends observed when comparing EOPD with controls. The enrichment was significant for CNVs overlapping PD-associated genes, including *PRKN*, and remained consistent when the analysis was restricted to exonic CNVs (Supplementary Figure 3).

### Survival analysis

Kaplan–Meier analysis revealed that individuals with a CNV in PD-related genes had significantly earlier symptom onset compared to those carrying CNVs in other genes or non-carriers (log-rank test, p-value = 7.0 e-06; Fig. 4B). Additionally, a Cox proportional hazards regression, adjusted for sex, and the first five ancestry PCs, indicated that having a CNV in PD-related genes was associated with a significantly increased hazard of earlier AAO (HR=1.48 [0.1-4.8]; *p-value* = 1.2e-06, Fig 4B). We further evaluated AAO exclusively in PD patients, comparing those with a CNV in PD-related genes to patients with other or no CNVs. The analysis also indicated that carrying at least one CNV leads to an earlier onset of symptoms (HR=1.39 [0.1-4.0]; p-value = 4.4e-05, Fig 4C).

## Discussion

CNVs have long been established as a major contributor to disease risk in a wide range of complex diseases including epilepsy, neuropsychiatric and neurodegenerative disorders ^10,22,29,30^. However, no large-scale study has been conducted at the genome-wide level to decipher the role of CNVs in sporadic PD. Thus, the current study aimed to extend the knowledge of potential role and impact of CNVs on PD by leveraging a well-characterized multinational PD cohorts from the COURAGE-PD and as well as the GP2 consortium. Given the lack of consensus on optimal methods for detecting and analysing genome-wide significant CNVs ^9^, we employed the recently developed sliding window method to identify potential novel significant CNV regions for PD using the sliding window approach ^34^. Such an approach has already been used to identify seizure-associated loci ^30^.

The use of array-based approaches to identify CNV-based loci can potentially lead to false-positive signals, thus, a careful approach to mitigate the risk of false positive findings is pertinent. Indeed, using two-stage design, our exploratory data set (COURAGE-PD) led to the identification of novel CNV regions including one deletion region and 13 duplication regions, which were associated with PD risk. The validation in an independent data set (GP2 consortium) failed to confirm the initial findings. This has been further confirmed by molecular validation using MLPA in individuals carrying CNVs within three of the most enriched CNV regions. These analyses did not confirm the presence of CNVs in the tested samples.

To explore the potential sources of this discrepancy, we undertook several methodological checks. A detailed inspection of CNV carrier distribution revealed a pronounced overrepresentation of samples from a single sub-cohort, across the genome-wide significant regions. This imbalance was not apparent through standard adjustments using principal components and cohort indicators. Instead, it became evident only after conducting leave-one-out GWAS analyses, in which the analyses demonstrated that the strongest association signals were primarily driven by samples from this particular sub-cohort. Interestingly, the individuals selected for MLPA validation were also drawn from this sub-cohort, which may explain the lack of CNV confirmation. When genome-wide significant CNV carriers were excluded, or when removing the entire sub-cohort was removed, all genome-wide significant signals disappeared.

PennCNV has long been the standard tool for CNV detection from SNP array data, particularly in large-scale studies. However, it frequently produces a high number of false positives, often exceeding 50% ^39^, despite the implementation of rigorous quality control procedures. This reflects the algorithm’s high sensitivity but limited specificity, a limitation further influenced by variables such as sample quality, array platform, as well as the number and density of SNPs within a genotyping array. Indeed, the initial findings from our study underscores this and cohort-specific artefacts and overrepresentation within one sub-cohort can easily be overlooked when employing standard pipeline for CNV discovery.

To address these challenges, various complementary strategies have been adopted, including the exclusion of low-quality samples and CNVs, cross-validation with alternative algorithms, and the application of stringent quality metrics ^40,41^. Nevertheless, given the potential impact of false discoveries, particularly when analyzing rare or pathogenic CNVs, functional validation of novel hits, using methods such as MLPA or qPCR remains essential for confirming CNV calls with high confidence.

Denser genotyping arrays tend to improve CNV detection by providing finer resolution and more accurate breakpoint definition, whereas arrays with sparse SNP coverage are more susceptible to technical artefacts and imprecise calls ^42–44^. In this study, CNVs were called using NeuroChip genotyping data, which includes approximately 300,000 common backbone SNPs, supplemented with rare variants targeting neurodegenerative disease loci. While the NeuroChip was specifically designed to enrich for known risk variants, its relatively low and uneven genome-wide SNP density may limit the accuracy of CNV detection, particularly in intergenic or gene-poor regions. This limitation could increase the risk of false positive calls and hinder precise breakpoint localization. In contrast, standard high-density genotyping arrays, such as the Illumina Global Screening Array or OmniExpress, typically include 600,000 to over 1 million SNPs, offering broader and more uniform genomic coverage that supports more robust structural variant detection. To evaluate whether SNP density and array content were primary contributors to technical noise in our CNV analysis, we repeated the GWAS using only the common variant backbone, thereby excluding most rare, disease-specific markers and approximating a more uniform genome-wide distribution. This analysis yielded results that were largely consistent with the initial findings, suggesting that the observed artefacts were not solely attributable to SNP number or density, but rather reflect broader limitations inherent to array-based CNV calling.

Our findings underscore these concerns, highlighting a critical limitation of array-based CNV detection: its vulnerability to technical artefacts that are not fully addressed by conventional covariate adjustments. Notably, associations identified in our discovery dataset failed to replicate in independent cohorts or through functional validation, emphasizing the need for stringent cohort-level quality control and replication strategies. These observations caution against overinterpretation of CNV signals driven by specific cohorts, and reinforce the importance of considering array design, including SNP density and genomic coverage, when evaluating CNV burden and conducting association analyses.

The AAO of PD is a key determinant of the clinical heterogeneity observed among patients. ^7,45,46^. In our study, we identified and validated a genome-wide significant deletion spanning the *PRKN* gene, which is well established as being associated with early-onset PD (EOPD). This finding aligns with the results of our burden analysis. *PRKN* homozygous or compound heterozygous deletions and duplications were identified as the most prevalent cause of EOPD and familial forms of PD ^47–49^. As compared to our CNV-based GWAS analyses, we did not identify any significant differences in the overall genome-wide CNV burden, similar to the previously published study ^22^. An increased burden of CNVs overlapping PD-related genes was observed in PD patients, primarily attributed to CNVs within the *PRKN* gene. This burden was even more pronounced in EOPD patients compared to controls and also to LOPD patients. Our observation is aligned with the published literature, in which it was shown that deletions and duplications accounted for 43.6% of all variants ^14^. The presence of these variants was found to be significantly associated with an earlier onset of PD. Furthermore, PD patients with a CNV in PD-related genes manifested a significantly earlier onset of symptoms compared to those with CNVs in other genes or non-carriers.

The majority of CNVs identified in the *PRKN* gene were heterozygous, but the presence of another pathogenic variant on the other allele cannot be ruled out. Heterozygous loss of *PRKN* function may elevate the risk of developing PD ^50–52^ and lead to an earlier AAO ^53^. However, the impact of heterozygous *PRKN* CNVs on PD susceptibility remains controversial. Some studies have indicated that carrying a heterozygous *PRKN* CNV may contribute to the development of PD, potentially through haploinsufficiency ^51^. Nevertheless, a study in subjects of European ancestry demonstrated no association between heterozygous *PRKN* CNVs and EOPD ^54^.

Power estimation for CNV-based studies remains an important area for future exploration, particularly given the potential of CNVs to contribute to disease risk. In our study, we identified a genome-wide significant deletion spanning the *PRKN* gene, well known to be involved in EOPD, highlighting the utility of our cohort and CNV detection pipeline in recovering biologically meaningful signals. However, several important limitations must be acknowledged. Most of the additional CNV associations did not replicate in independent cohorts and functional validation, suggesting they were likely artifacts driven by technical noise or cohort-specific biases. Although CNVs were called using widely accepted best practices applied to large-scale genotyping data, the reliance on genotyped SNPs for breakpoint estimation inherently limits resolution. This issue is particularly relevant in our dataset where the limited SNP density likely contributed to both false positives and imprecise breakpoint definition. While MLPA or qPCR were not applied to all CNVs in our study, prior validations of *PRKN* and *SNCA* CNVs in PD cohorts have demonstrated high confirmation rates, supporting the reliability of these specific findings. To partially address the resolution limitation, we applied a sliding window approach to identify recurrent regions of CNV burden. Nevertheless, CNV-GWASs continue to face unique challenges, particularly in pinpointing the causal gene(s) within large genomic intervals affected by CNVs. Future studies using higher-resolution platforms, such as whole-genome sequencing, and involving more ethnically diverse populations will be essential to clarify the contribution of rare CNVs to PD risk.

## Data Availability

Genome-wide CNV association summary statistics are available in the Supplementary Data. Individual-level CNVs and genotyping data are available on request from the COURAGE-PD consortium. Relevant scripts used in the present work are available on GitHub (https://gitlab.lcsb.uni.lu/genomeanalysis/cnv_gwas_courage-pd). For the sliding window association method, we used the code available under the Talkowski Laboratory (Massachusetts General Hospital & The Broad Institute) repository in Zenodo (https://github.com/talkowski-lab/rCNV2/tree/v1.0, with https://zenodo.org/records/6647918).

## Acknowledgments

This study used data from the Courage-PD consortium, conducted under a partnership agreement among 35 studies. The Courage-PD consortium is supported by the EU Joint Program for Neurodegenerative Disease research (JPND https://neurodegenerationresearch.eu). This project was supported by the Global Parkinson’s Genetics Program (GP2; https://gp2.org). GP2 is funded by the Aligning Science Across Parkinson’s (ASAP) initiative and implemented by The Michael J. Fox Foundation for Parkinson’s Research (MJFF). The complete list of GP2 members is provided supplementary table 5. P. May was funded by the Fonds National de Recherche (FNR), Luxembourg, as part of the National Centre of Excellence in Research on Parkinson’s Disease (NCER-PD, FNR11264123). Z. Landoulsi and P. May were supported by the DFG Research Unit FOR2715 (INTER/DFG/17/ 11583046), FOR2488 (INTER/DFG/19/14429377) and the National Centre for Excellence in Research on Parkinson’s disease (NCER-PD). A.B. Singleton, D.G. Hernandez, and C. Edsall are funded by the Intramural Research Program of the National Institute on Aging, National Institutes of Health, Department of Health and Human Services, project ZO1 AG000949. E. Rogaeva is funded by the Canadian Consortium on Neurodegeneration in Aging. S.Koks is funded by MSWA.

P. Taba is the recipient of an Estonian Research Council Grant PRG957. E.M.Valente is funded by the Italian Ministry of Health (Ricerca Corrente 2021). S. Bardien and J. Carr are supported by grants from the National Research Foundation of South Africa (grant number: 106052); the South African Medical Research Council (Self-Initiated Research Grant); and Stellenbosch University, South Africa; they also acknowledge the support of the NRF-DST Centre of Excellence for Biomedical Tuberculosis Research; South African Medical Research Council Centre for Tuberculosis Research; and Division of Molecular Biology and Human Genetics, Faculty of Medicine and Health Sciences, Stellenbosch University, Cape Town. P. Pastor have received funding from the Spanish Ministry of Science and Innovation (SAF2013-47939-R). K. Wirdefeldt and N.L. Pedersen are funded by the Swedish Research Council, grant numbers K2002-27X-14056-02B, 521-2010-2479, 521-2013-2488, and 2017-02175. N.L. Pedersen is funded by the National Institutes of Health, grant numbers ES10758 and AG 08724. C. Ran is funded by the Märta Lundkvist Foundation, Swedish Brain Foundation, and Karolinska Institutet Research Fund. A.C. Belin is funded by the Swedish Brain Foundation, Swedish Research Council, and Karolinska Institutet Research Funds. M. Tan is funded by the Parkinson’s UK. M. Sharma was supported by grants from the German Research Council (DFG/SH 599/6-1), MSA Coalition, and The Michael J. Fox Foundation (USA Genetic Diversity in PD Program: GAP-India Grant ID: 17473). A. Elbaz reports grants from Agence nationale de recherche (ANR), The Michael J. Fox Foundation, Plan Ecophyto (French Ministry of Agriculture), and France Parkinson outside the submitted work. PG GEN sample collection was funded by the MRC and UK Medical Research Council (C.E. Clarke and K.E. Morrison). The sponsors had no role in the study design, data collection, data analysis, data interpretation, writing of the report, or decision to submit the paper for publication.

## Author contributions

Z.L, P.M, M.S, RK contributed to the conception and design of the study. Z.L, A.A.K.S, N.K, C.S, D.R.B, L.M, C.L, L.M.N, E.H, C.D, L.Pavelka, P-E.S, M.R, P.L, B.P, C.E, J.K, D.G.H, C.B, G.D.M, A.Zimprich, W.P, M.Tan, E.R, A.L, S.K, P.T, S.L, A.B, J-C.C, M-C.C-H, E.M, K.B, A.B.D, G.M.H, E.D, L.Stefanis, A.M.S, D.V, E.M.V, S.P, L.Straniero, A.Zecchinelli, G.P, L.B, C.F, G.A, A.Q, M.G, L.F.B, H.M, A.N, N.H, K.N, S.J.C, Y.J.K, P.K, B.P.C.V.D.W, B.R.B, A.B.S, M.Toft, L.Pihlstrom, L.C.G, J.J.F, S.B, J.C, E.T, M.E, P.P, K.W, N.L.P, C.R, A.C.B, A.P, C.E.C, K.E.M, D.K, M.J.F, A.E, T.G, D.L contributed to the acquisition and analysis of data. ZL, PM, MS, RK contributed to drafting the text or preparing the figures.

All the authors contributed to revise the manuscript and approved the submitted version.

