## Supplementary material for "Genome-wide association study of copy number variations in Parkinson’s disease": Supplemetal data

**Supplementary Tables**

**Table S1: Characteristics of PD and controls from the COURAGE-PD consortium by study site after quality control.**

**Table S2: Summary of CNV calls after quality control steps in PD and controls.**

**Table S3: Summary of CNV calls in PD-related genes.**

**Table S4: CNVs in PD-related genes (See excel sheet Table_S4).**

**Table S5: List of GP2 members (GP2_authors_affiliations.pdf)**

**Supplementary Figures**

**Figure S1: Genome-wide meta-analysis of PD in Tübingen cohort.**

**Figure S2: Genome-wide meta-analysis of PD using CNVs called only in the NeuroChip backbone.**

**Figure S3: CNV burden in early-onset vs late-onset PD patients**

**Supplementary Tables**

**Table S1: Characteristics of PD and controls from the COURAGE-PD consortium by study site after quality control.**

| **study** | **Country** | **Status** | **N** | **Age at assessment** | **Sex** | | **Age at onset** |
| --- | --- | --- | --- | --- | --- | --- | --- |
|  |  |  |  | **Mean (SD, MD)** | **% Male** | **% Female** | **Mean (SD, MD)** |
| Aasly | Norway | PD | 510 | 77.7 (11.6, 79) | 60 | 40 | 60.5 (11, 0) |
|  |  | Controls | 509 | 73 (14.1, 0) | 54.9 | 45.1 |  |
| Annesi | Italy | PD | 93 | 66.5 (9, 0) | 64.6 | 35.4 | 59.2 (10.1, 0) |
|  |  | Controls | 94 | 58.2 (20.3, 0) | 45.8 | 54.2 |  |
| Bardien/Carr | South Africa | PD | 170 | 68.7 (10.6, 0) | 61.2 | 38.8 | 60 (11.9, 0) |
|  |  | Controls | 85 | 51.1 (12.7, 0) | 56.5 | 43.5 |  |
| Brice/Corvol/Lesage | France | PD | 851 | 60.7 (10.9, 32) | 58.5 | 41.5 | 51.9 (11.1, 32) |
|  |  | Controls | 280 | 62.4 (10.7, 0) | 56.5 | 43.5 |  |
| Carmine Belin/Ran | Sweden | PD | 286 | 67.4 (10.3, 0) | 61.6 | 38.4 | 59.4 (11.1, 0) |
|  |  | Controls | 629 | 66.5 (10.4, 597) | 55.2 | 44.8 |  |
| Chartier-Harlin/Mutez | France | PD | 370 | 64.3 (9, 0) | 54.4 | 45.6 | 52.7 (10.5, 0) |
|  |  | Controls | 228 | 59.9 (12.6, 0) | 40 | 60 |  |
| Deutschländer | Germany | PD | 285 | 69.4 (9.9, 0) | 61.8 | 38.2 | 60.4 (11.5, 0) |
|  |  | Controls | 46 | 66 (10.3, 0) | 30.5 | 69.5 |  |
| Elbaz | France | PD | 455 | 70.1 (7.4, 0) | 59.8 | 40.2 | 64.9 (7.7, 0) |
|  |  | Controls | 1066 | 69.8 (7.6, 0) | 59.2 | 40.8 |  |
| Farrer | United States | PD | 389 | 67.7 (10.3, 1) | 65.6 | 34.4 | 56.5 (11.1, 1) |
|  |  | Controls | 409 | 69.4 (12.4, 0) | 31.8 | 68.2 |  |
| Ferreira | Portugal | PD | 415 | 69 (10.1, 0) | 57.2 | 42.8 | 57.9 (11.9, 0) |
|  |  | Controls | 67 | 47.1 (17.8, 0) | 25.4 | 74.6 |  |
| Gasser/Sharma | Germany | PD | 1664 | 65.1 (10.3, 0) | 62.8 | 37.2 | 59.6 (10.9, 0) |
|  |  | Controls | 1070 | 60.7 (7.4, 0) | 50.8 | 49.2 |  |
| Duga/Cilia | Italy | PD | 1391 | 65.8 (10.8, 0) | 59.3 | 40.7 | 59.1 (11.2, 0) |
|  |  | Controls | 1370 | 61.9 (10.9, 0) | 33.8 | 66.2 |  |
| Hadjigeorgiou | Greece | PD | 283 | 67.9 (10.3, 0) | 48.5 | 51.5 | 63.1 (10.1, 0) |
|  |  | Controls | 314 | 69.8 (8.7, 0) | 47.2 | 52.8 |  |
| Koks/Taba | Estonia | PD | 216 | 73 (8.2, 0) | 39.9 | 60.1 | 66.8 (9.8, 0) |
|  |  | Controls | 170 | 72.3 (10.2, 0) | 41.8 | 58.2 |  |
| Kruger | Luxembourg | PD | 333 | 67.4 (11.2, 0) | 67.3 | 32.7 | 59.7 (13, 0) |
|  |  | Controls | 360 | 58.1 (11.9, 0) | 55.9 | 44.1 |  |
| Mellick | Australia | PD | 480 | 68.3 (9.3, 0) | 64 | 36 | 59.1 (11.3, 0) |
|  |  | Controls | 508 | 66.9 (9.6, 0) | 45.3 | 54.7 |  |
| Pastor/Diez-Fairen | Spain | PD | 499 | 67 (10.6, 2) | 57.2 | 42.8 | 58.9 (11.9, 2) |
|  |  | Controls | 338 | 66 (11.3, 0) | 40.9 | 59.1 |  |
| Puschmann | Sweden | PD | 110 | 68.8 (10.6, 0) | 62.8 | 37.2 | 60.5 (10.6, 0) |
|  |  | Controls | 105 | 66.7 (8.9, 0) | 29.6 | 70.4 |  |
| Rogaeva/Lang | Canada | PD | 215 | 63.6 (12.4, 6) | 66.1 | 33.9 | 54 (12.2, 6) |
|  |  | Controls | 153 | 73.8 (8.6, 0) | 38 | 62 |  |
| Stefanis/Simitsi | Greece | PD | 242 | 67.1 (13.5, 0) | 61.6 | 38.4 | 60.7 (13.6, 0) |
|  |  | Controls | 181 | 67 (9.6, 0) | 36 | 64 |  |
| Toft | Norway | PD | 480 | 65.7 (8.9, 0) | 63 | 37 | 55.9 (11.3, 0) |
|  |  | Controls | 442 | 62 (11.1, 0) | 55.7 | 44.3 |  |
| Tolosa | Spain | PD | 309 | 66 (10.6, 0) | 60.6 | 39.4 | 58.5 (11.3, 0) |
|  |  | Controls | 68 | 62.3 (12.1, 8) | 16.2 | 83.8 |  |
| Valente | Italy | PD | 326 | 67 (12.1, 7) | 62.3 | 37.7 | 55.3 (10.9, 7) |
|  |  | Controls | 54 | 78.4 (9.6, 0) | 38.9 | 61.1 |  |
| Wirdefeldt | Sweden | PD | 65 | 75.7 (8.1, 0) | 53.9 | 46.1 | 66.6 (9.9, 0) |
|  |  | Controls | 171 | 73.6 (9.8, 0) | 45.7 | 54.3 |  |
| Zimprich | Austria | PD | 598 | 66.8 (11.3, 0) | 63.4 | 36.6 | 58.8 (11.3, 0) |
|  |  | Controls | 184 | 0 (0, 184) | 40.8 | 59.2 |  |

**Table S2: Summary of CNV calls after quality control steps in PD and controls.**

|  | **Controls** | **PD** |
| --- | --- | --- |
| **Number of samples** | 8901 | 11035 |
| **CNV carriers (n, %)** | 3299 (38.7 %) | 3896 (35.8 %) |
| **Number of CNVs** | 15544 | 12719 |
| **Duplication** | 2822 | 5021 |
| **Deletions** | 12722 | 7698 |
| **CNVs per sample (Mean, SD)** | 4.71 (9.9) | 3.26 (9.5) |
| **Mean size of CNVs (Kb, Mean, SD)** | 241 (1693) | 266 (1766) |
| **Number of SNPs per CNV (Mean, SD)** | 126 (578) | 149 (552) |
| **filtered out CNVs** | 551815 | 518143 |
| **filtered out duplication** | 160046 | 228843 |
| **filtered out deletions** | 391769 | 289300 |

**Table S3: Summary of CNV calls in PD-related genes.**

| **Gene** | **PD (N = 151)** | | **Controls (N = 78)** | |
| --- | --- | --- | --- | --- |
|  | 73 del | 78 dup | 28 del | 50 dup |
| ***LRRK2*** | 0 | 3 | 0 | 0 |
| ***PARK7*** | 1 | 0 | 0 | 0 |
| ***PRKN*** | 71  (7 hmz, 7 intronic) | 64  (29 intronic) | 28  (5 intronic) | 47  (17 intronic) |
| ***SNCA*** | 1 | 7 | 0 | 1 |
| ***VPS35*** | 0 | 4 | 0 | 2 |

**Table S4: CNVs in PD-related genes (**See Excel sheet “Table_S4”)

CNV coordinates column (chr:start-end), Copy number (0: two-copies deletions, 1: one copy deletion and 3 one copy duplication), No Probes (number of SNPs overlapping the CNV), Samples were anonymized with two codes PD for patients and CTL for controls. Validation by MLPA: * CNV validated and ** CNV not validated.

**Supplementary Figures**

**Figure S1: Genome-wide meta-analysis of PD in Tübingen cohort.** Miami plot of the CNV genome-wide association analyses illustrating the -log10 transformed Bonferroni-corrected p-values (DEL and DUP for deletions at the top and duplications at the bottom, mirrored respectively) for the enrichment of CNVs in cases vs. controls for each 200 kb sliding window. Genomic regions that exceeded the Bonferroni-corrected significance threshold (blue line, α =3.74×10^-6^) were annotated with the genomic band containing the signal.

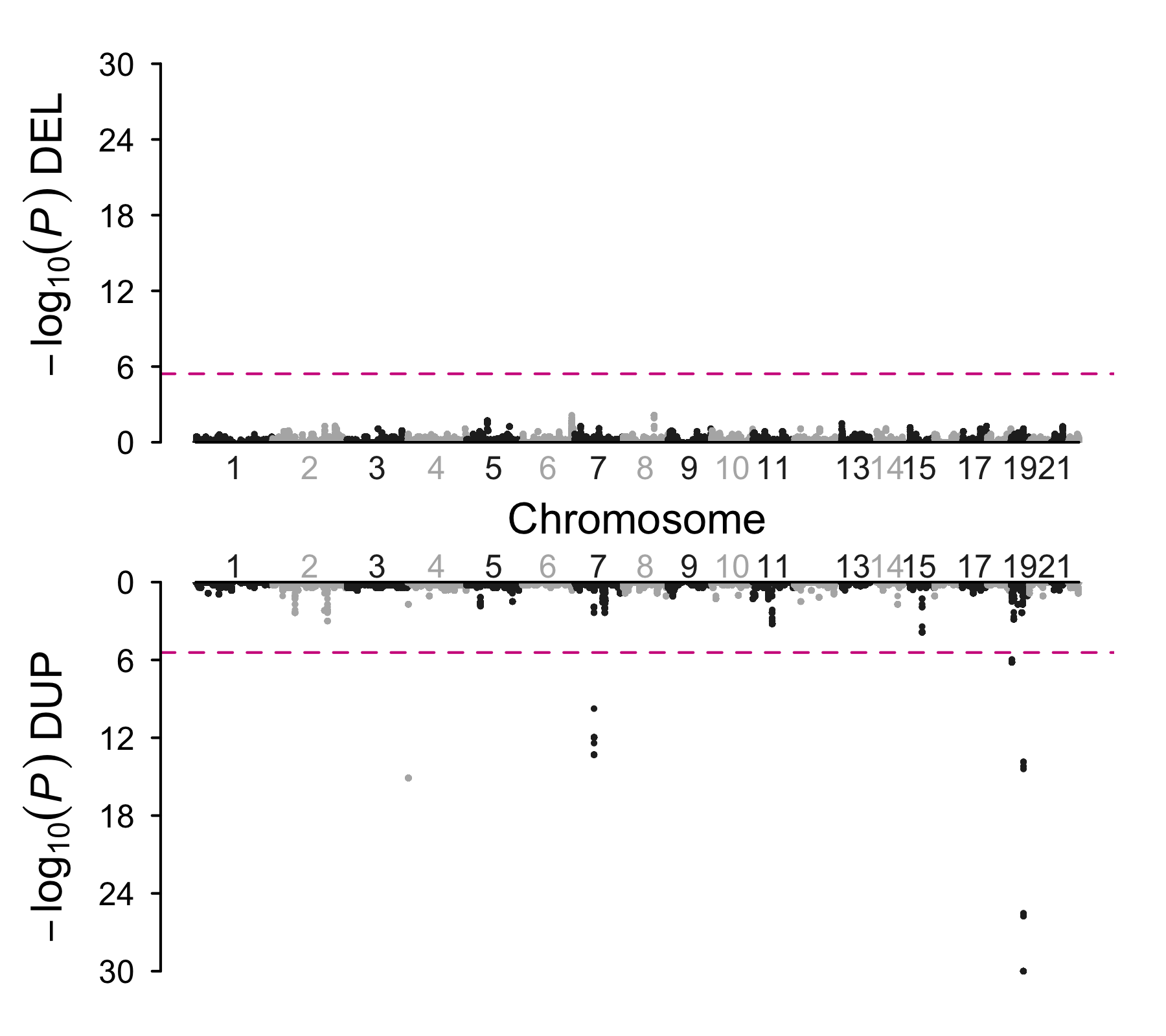

**Figure S2: Genome-wide meta-analysis of PD using CNVs called only in the NeuroChip backbone.** Miami plot of the CNV genome-wide association analyses illustrating the -log10 transformed Bonferroni-corrected p-values (DEL and DUP for deletions at the top and duplications at the bottom, mirrored respectively) of the Fisher’s exact tests for the enrichment of CNVs in cases vs. controls for each 200 kb sliding window before (A) and after (B) the exclusion of 220 samples from the Tübingen cohort carrying genome-wide significant CNVs. Adjacent chromosomes are shown in alternating light and dark colors. Genomic regions that exceeded the Bonferroni-corrected significance threshold (blue line, α =3.74×10^-6^) were annotated with the genomic band containing the signal.

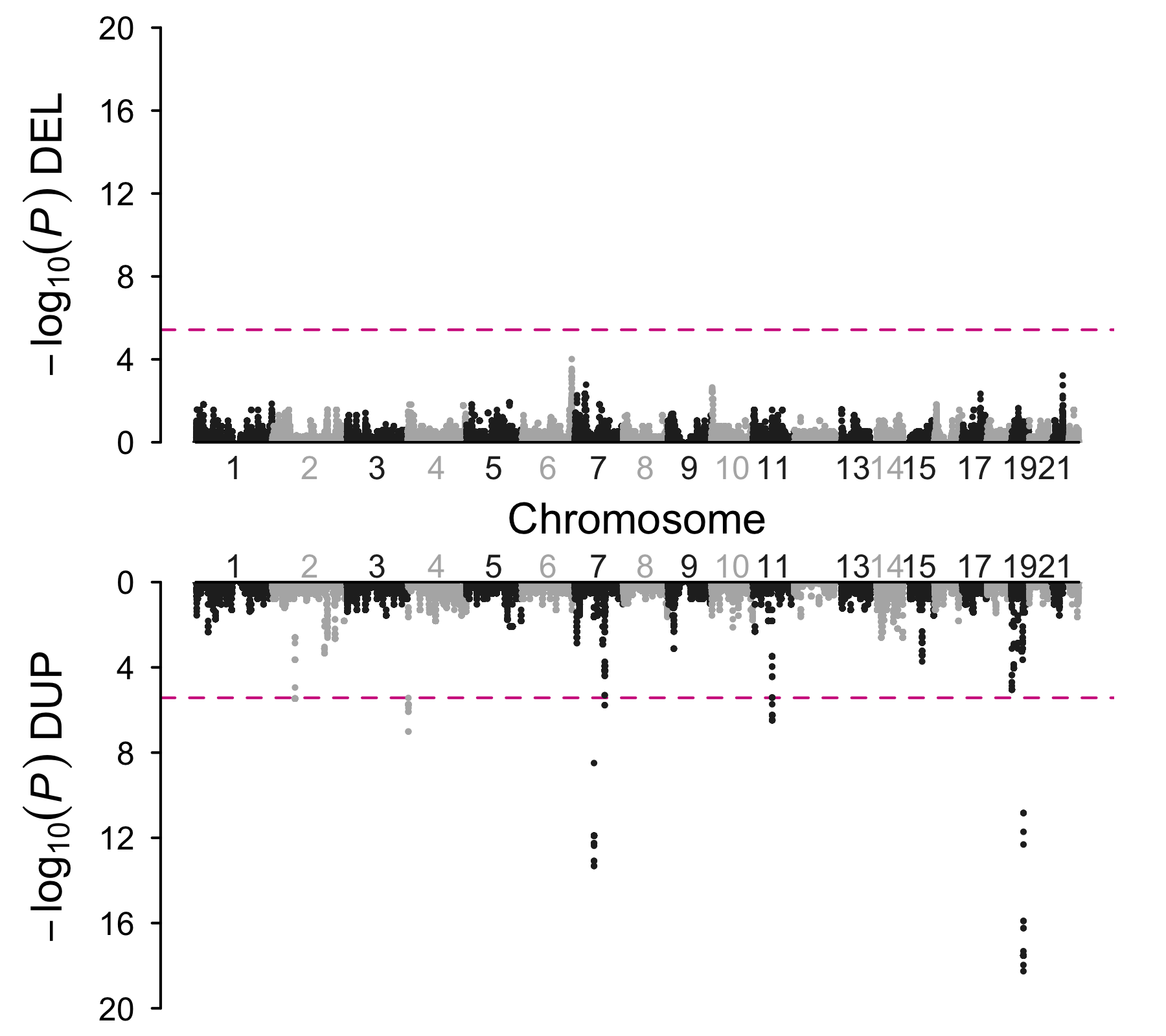

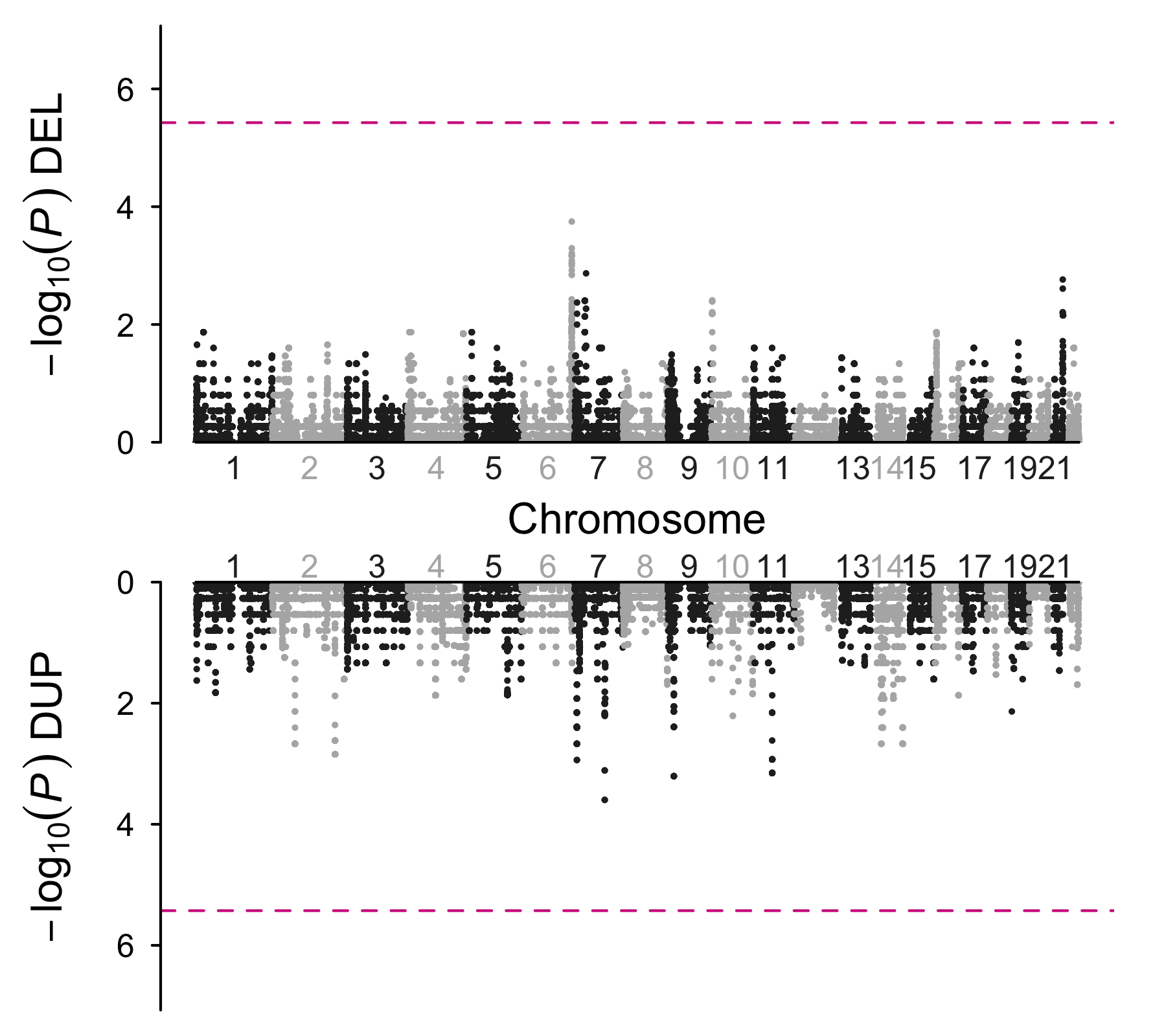

**A**

**B**

**Figure S3: CNV burden in early-onset vs late-onset PD patients.** Logistic regression was used to calculate odds ratios (ORs) and Bonferroni-adjusted p-values for each CNV category, and were adjusted for age, sex, and the first five components of PCAs. Protein-coding gene categories were defined as all coding genes except PD-related genes. * Bonferroni adjusted p-values surpassing the multiple testing cut-off.

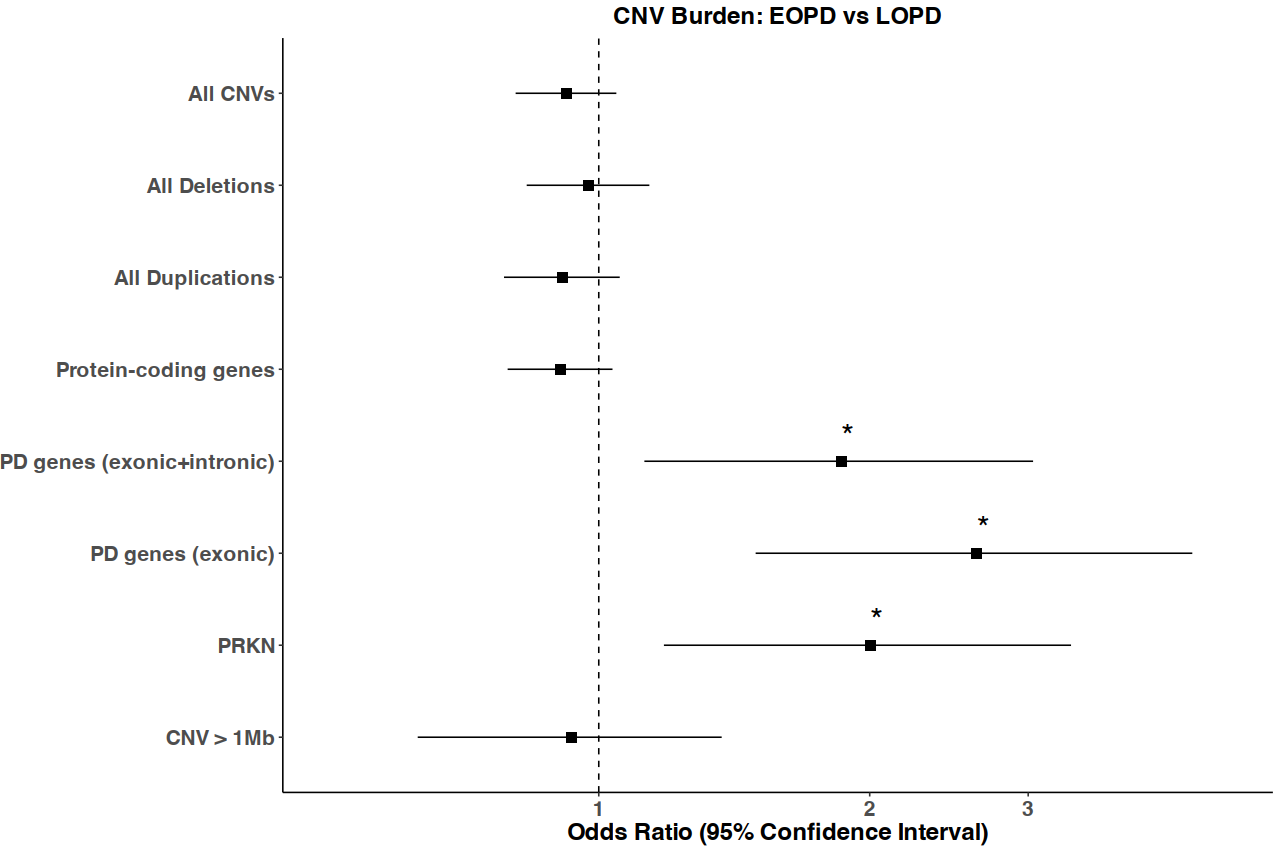
